# Consequences of Acute Presentations of Functional Neurological Disorders in Neuro-oncology patients: Case Series and Systematic Review

**DOI:** 10.1101/2024.11.14.24317315

**Authors:** Stuart C. Innes, Dorothy K. Joe, Katia Cikurel, José P. Lavrador, Francesco Vergani, Ranjeev Bhangoo, Keyoumars Ashkan, Gerald T. Finnerty

## Abstract

**Objective:** New neurological symptoms in neuro-oncology patients are usually attributed to the tumour or its treatment. A diagnosis of functional neurological disorder (FND) is often only considered when investigations do not reveal a cause and medical management fails. Here, we investigate the consequences of acute FND in neuro-oncology patients.

**Methods:** We performed a single-centre retrospective case study of adult neuro-oncology patients with an intracranial tumour who presented acutely with FND. Data recorded were: tumour type, investigations, adverse life events, medical interventions and outcomes. This was combined with a systematic literature review of articles in English peer-reviewed journals on adults with brain or meningeal tumours and concomitant FND.

**Results:** Ten patients met our study criteria. Six had functional seizures, two had functional hemiparesis and two had functional speech disorders. FND symptoms started: prior to tumour diagnosis in three patients; between diagnosis and treatment in four; and after treatment commenced in three patients. Two patients were thrombolysed for presumed stroke. Three patients had their tumour surgery or chemoradiotherapy delayed. Diagnosis and management of FND enabled tumour treatment to restart. The systematic review identified 37 patients. 33 had functional seizures and four had motor FND. All FND diagnoses except one started after tumour treatment commenced.

**Conclusion:** Acute FND may lead to unnecessary medical interventions and disrupt tumour treatment. Although acute FND improved with diagnosis and explanation, many neuro-oncology patients require a multi-disciplinary tumour-FND pathway to manage their FND, prevent acute FND symptoms becoming chronic and to avoid delays to tumour treatment.

**What is already known on this topic:** Case reports have documented functional neurological disorders (FND) in brain tumour patients, but almost invariably after their tumour treatment has commenced.

**What this study adds:** FND can present acutely at any stage of the brain tumour illness and may result in inappropriate interventions and/or treatment. Tumour treatment may be delayed.

**How this study might affect research, practice or policy:** Brain tumour patients require a multi-disciplinary FND pathway, not only to manage their FND symptoms, but to prevent delays to tumour treatment, which is proscribed by government guidance on timely treatment of cancer.

## INTRODUCTION

People with brain and meningeal tumours report a wide variety of symptoms, such as seizures, focal neurological deficits, headache and cognitive dysfunction.^1 2^ The symptoms usually reflect the tumour location or, later on, may be side effects from tumour treatment.^3 4^ There is increasing awareness that a functional neurological disorder (FND) can cause new symptoms in people with neurological conditions not due to tumours.^5 6^ Little is known about the extent to which FND can cause new symptoms in people with brain or meningeal tumours and the consequences of those FND symptoms for tumour management.

Commonly, new neurological symptoms in patients with brain tumours are initially attributed to the tumour or its treatment. A diagnosis of FND is often only considered when investigations are inconclusive and medical management fails rather than being made using positive, “rule-in” clinical signs of FND.^7 8^

People with FND, including those with brain tumours, may present acutely to the local Emergency department and be admitted for investigation.^9^ A concern is that these admissions may result in inappropriate medical interventions^10^ or treatment^11^.

The consequences of acute FND in people with brain and meningeal tumours has received little attention. In particular, the ramifications of acute FND on tumour management have not been explored. To address these issues, we focused on brain tumour patients who presented acutely with FND that required expedited investigations. Commonly, this was because the FND symptoms resembled complications associated with the tumour that required prompt action, e.g. tumour progression; major side effects of tumour treatment; or development of disabling, tumour-associated conditions, such as stroke. We complemented our case series with a systematic review of the literature on co-occurrence of FND in people with brain and meningeal tumours.

## MATERIALS AND METHODS

### Case Series

We performed a retrospective case review of adult patients (≥ 18 years old) attending a regional Neuro-oncology service from mid-2017 to mid-2021 who presented acutely with symptoms requiring expedited investigations which were later attributed to FND. The diagnosis of FND was made in accordance with the DSM-5 text revision^12^ by a Consultant Neurologist working in the Neuro-oncology service and was based on history, clinical examination emphasising positive signs of FND,^7 8^ investigation results and confirmed with patient follow-up.

Clinical data were obtained by reviewing all note types available within the electronic patient records of King’s College Hospital and included: history, clinical examination findings, brain tumour type, diagnostic investigations, psychiatric co-morbidities and adverse life events, therapeutic interventions for FND and outcomes.

### Systematic Review

A systematic literature review was performed to enable comparison of our study with the literature and to facilitate recommendations from the study. The systematic review was registered with PROSPERO 2021 CRD42021250037 and is available from: https://www.crd.york.ac.uk/prospero/display_record.php?ID=CRD42021250037) And performed between April 2017 and 2021 for presentation. An additional search was performed covering records from 2021 to May 2024.

Inclusion criteria were articles published in English peer-reviewed journals on adults (age ≥18 years) who had either brain tumours or meningeal tumours and, also, had FND. All study types were included regardless of size. We searched PubMed, EMBASE, PsychINFO, Web of Science and Cochrane Library databases for studies published in English between 1943 and April 2024 using the search strategy: (Brain neoplasms OR neuro-oncology OR glioma OR glioblastoma OR astrocytoma OR meningioma OR schwannoma OR oligodendroglioma OR medulloblastoma OR ependymoma OR brain tumour OR brain tumor OR brain cancer OR brain neoplasia) AND (“conversion disorder” OR psychogenic OR “non epileptic” OR hysteria OR “functional neurological disorder” OR “functional disorder” OR “functional movement” OR “functional motor” OR “somatization” OR “non-epileptic attack disorder” OR “NEAD” OR “PNES”). Articles were excluded if the full text could not be obtained or physical symptoms were attributed to an underlying psychiatric disorder, not FND. The last database searches were completed on 3 May 2024.

Articles identified from electronic searches were downloaded into reference management software. Duplicated articles were removed. The articles were screened independently by two reviewers (SCI, DKJ). If the Abstract met the study criteria, the full-text was scrutinized by both reviewers for the following information: study design; participant demographics; presenting functional neurological symptoms, onset of functional neurological symptoms with respect to tumour diagnosis and treatment, that is before the tumour was first diagnosed, after first diagnosis and before treatment had begun or after tumour treatment had begun; FND diagnostic investigations; time to diagnosis, presence/absence of psychiatric co-morbidities and significant adverse life events; and FND management and outcomes. Missing outcome data are reported in Table 2.

Primary outcomes were: clinical symptoms at presentation of the FND; time of FND diagnosis divided into: before tumour diagnosis; between tumour diagnosis and start of the first round of tumour treatment; and after first round of treatment had started. Secondary outcomes included any evidence of psychological co-morbidities or adverse life events prior to the neuro-oncology diagnosis. All identified articles were case reports and were characterized with the quality assessment tool by Murad et al.^13^ (Supplementary Table 1).

If the neuro-oncology lesion was managed conservatively, that is had no neurosurgical or radio-chemotherapy, then tumour management was deemed to have started at the time of diagnosis. Discrepancies over whether an article met the study criteria were resolved by a third reviewer (GTF).

## RESULTS

### Case series

We identified ten adult patients (median age, 46 years) who met the inclusion criteria (Table 1). Four patients had a lesion involving their right cerebral hemisphere and six patients had a lesion involving their left cerebral hemisphere.

**Table 1:**
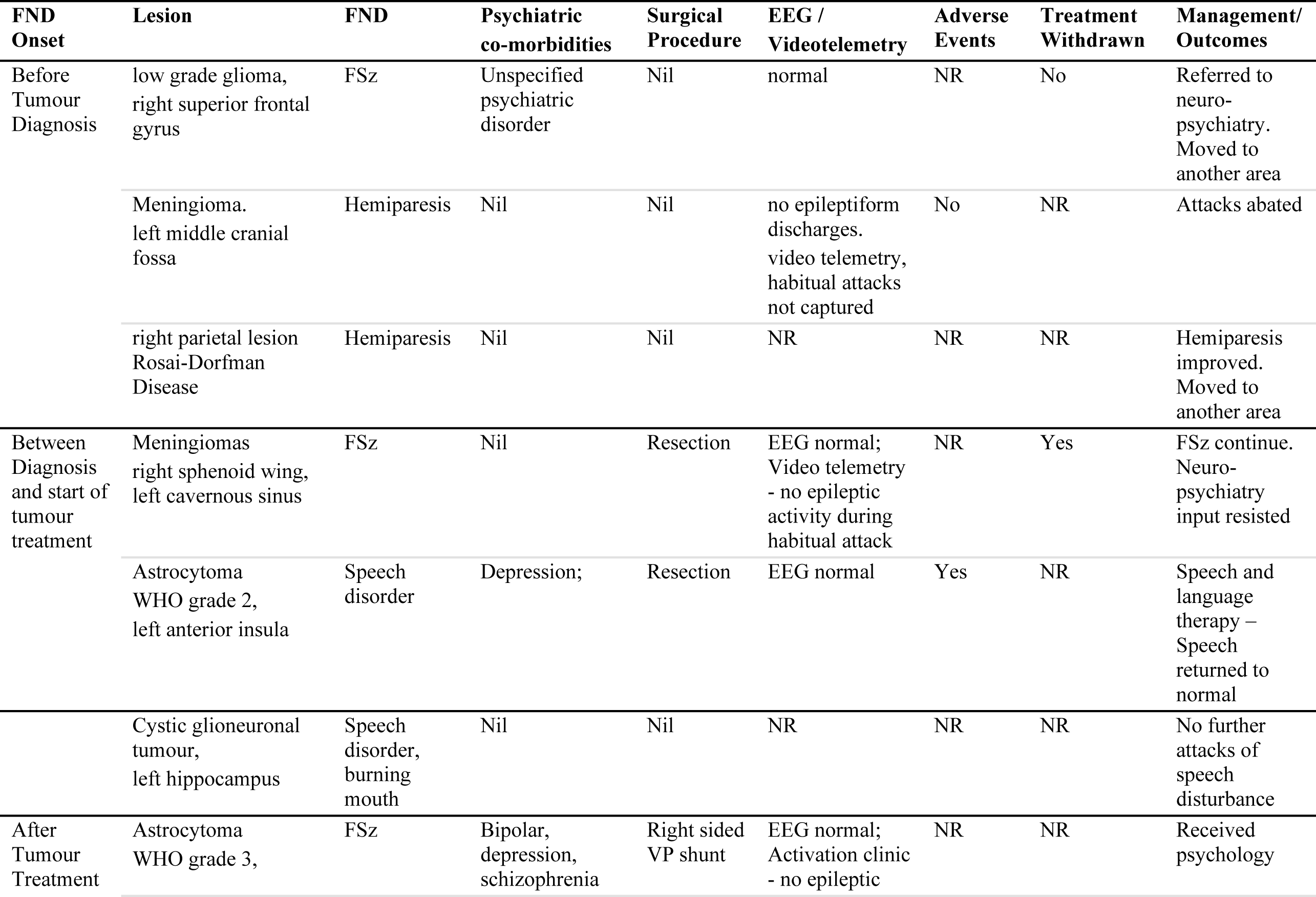
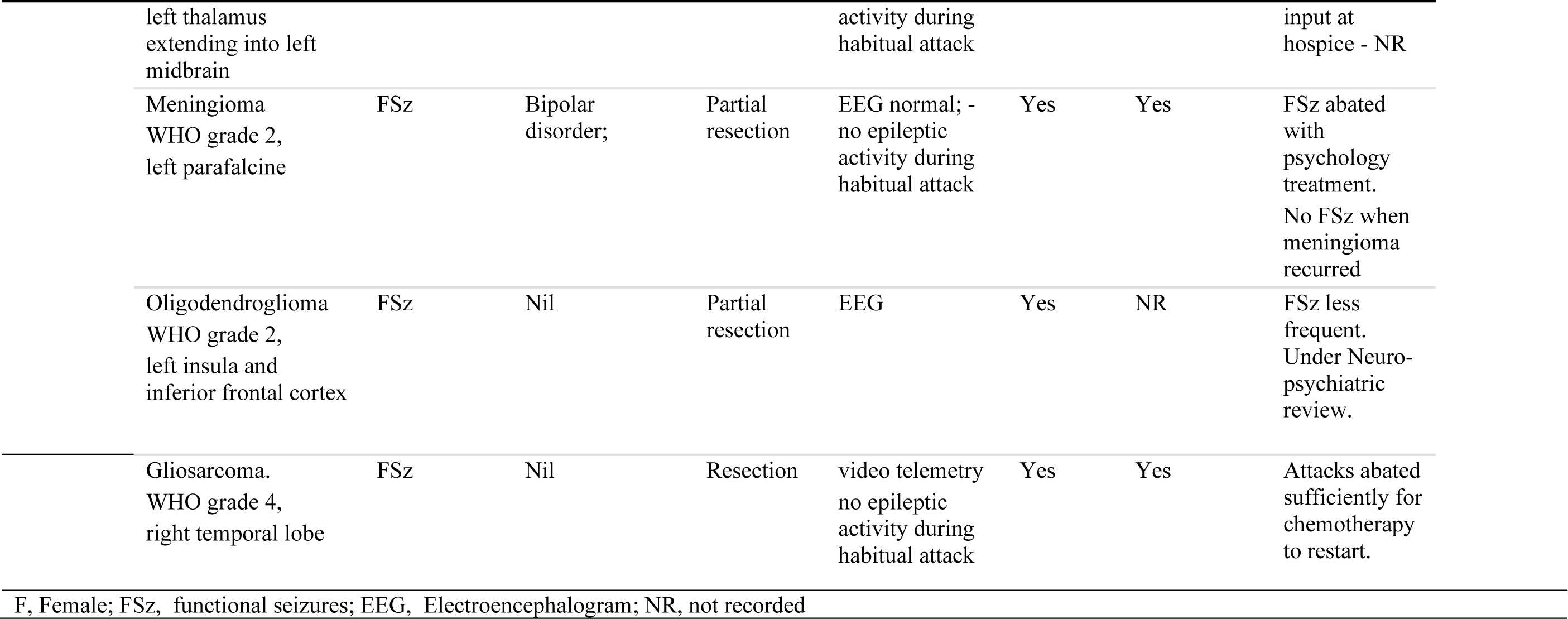
Neuro-oncology patients presenting acutely with FND.

#### Acute FND in neuro-oncology patients

Functional seizures (also known as psychogenic non-epileptic seizures, dissociative seizures or non-epileptic attack disorder)^14 15^ were the most common acute FND presentations, occurring in six patients. Hemiparesis developed in two patients (one right-sided, one left-sided). Two patients presented with a functional speech disorder, which manifested as either a new onset stutter or intermittent episodes of speech arrest combined with burning mouth syndrome (Table 1).

Tumour-associated seizures are a common symptom of brain tumours. Two of the six patients with functional seizures had tumour-associated seizures prior to the tumour diagnosis being established. The semiology of the functional seizures was different from the tumour-associated seizures in the patients that had both tumour-associated seizures and functional seizures. The remaining four patients had de novo functional seizures.

Seven of ten patients had either a lower-grade glioma (n = 5) or a meningioma WHO grade 1 (n = 2) (Table 1). The three remaining patients had either a gliosarcoma or a meningioma WHO grade 2 or Rosai-Dorfman disease.

#### Timing of acute FND

We investigated when acute FND presented in neuro-oncology patients. Seven of ten patients developed acute FND (functional seizures, 3; functional hemiparesis, 2; functional speech disorders, 2) in the interval between the start of their neurological investigations and 6 weeks after the start of their first round of tumour treatment.

We explored the timing of the acute FND further by dividing the tumour illness into three periods: before diagnosis, between diagnosis and tumour treatment starting and after treatment had started (Figure 1). Functional neurological symptom onset occurred before tumour diagnosis in three patients (30%). Two of these three patients developed a functional hemiparesis during the period when they were having neurological investigations leading up to a tumour diagnosis (Table 1). The remaining patient initially presented with functional seizures on the background of a psychiatric disorder and was later (>1 year) found to have a brain tumour. An acute FND developed in three patients (30%) in the interval between tumour diagnosis and treatment (functional seizure, 1; functional speech disorder, 2). The remaining four patients (40%) all developed functional seizures after treatment had started with varying delays (<1 week, 1; 6 weeks, 1; >1 year, 2). We concluded that acute FND could occur at any stage of the tumour-illness journey. However, most patients developed acute FND in the period between tumour investigations to six weeks after starting treatment.

**Figure 1.**
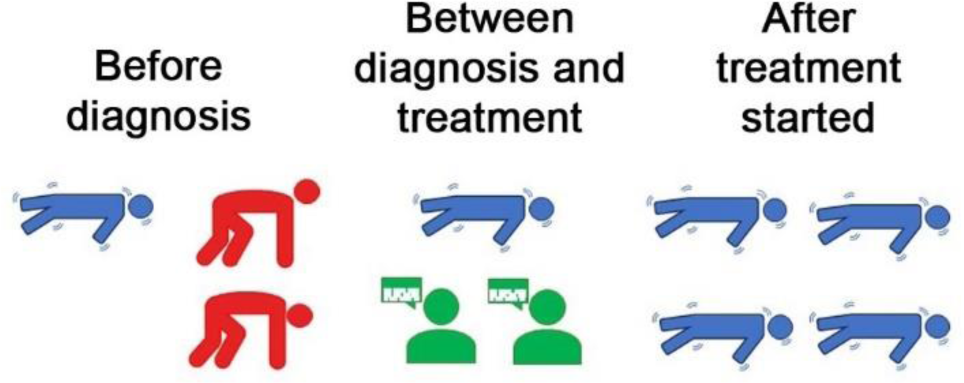
Timing of FND in relation to tumour diagnosis and treatment. The colour-coded people-icons denote: Blue, seizure; red, hemiparesis; green, speech disturbance Each patient is represented by one icon.

#### Risk factors for acute FND

We looked at possible risk factors for neuro-oncology patients developing an acute FND.^7 16 17^ Four patients (40%) had a psychiatric diagnosis before the tumour was diagnosed. Serious adverse life events (mental, physical, and sexual abuse; exposure to crime; close relative terminal illness or bereavement) were described by four patients. One patient reported no adverse life events. Information was not available for the remaining five patients.

#### Consequences of acute FND for neuro-oncology patients

We then examined the consequences for neuro-oncology patients of an acute FND. Two patients (one functional hemiparesis, one functional speech disorder) were initially diagnosed as strokes and received intravenous thrombolysis.

Diagnosis and management of acute FND symptoms using psychiatric, psychological and speech therapies was associated with progressive improvements to FND symptoms over several months in seven (70%) patients. The three patients who did not show improvement in their FND symptoms all had functional seizures and a comorbid psychiatric disorder.

Tumour treatment continued in parallel with management of the acute FND. However, neuro-oncology care was interrupted for at least one month in three patients (30%) by delaying neurosurgery in one patient (Example case history) or by disrupting subsequent oncology treatment in two patients. Tumour treatment restarted when the FND symptoms had been explained to the patient and the FND symptoms had either abated (Example case history) or had improved to the point that they could be managed in parallel with tumour treatment. Precise measurement of the treatment delay was hampered by the COVID-19 pandemic, which prolonged treatment delays in some patients.

Six patients who presented with functional seizures were on high doses of anticonvulsant medication. The dose of anticonvulsant medication was markedly reduced or stopped in three of those patients (3/6, 50%) after functional seizures were diagnosed.

We explored the longer-lasting consequences of acute FND by reviewing patients’ follow-up over the 4-year study period. One patient died one year after presenting with functional seizures and three patients (functional seizures, 2; functional hemiparesis, 1) left the area. At the time of leaving the area, the functional seizures in the two affected patients had not improved whereas the functional hemiparesis had resolved. Of the six remaining patients, three (2 functional speech disorder, 1 functional seizures) have had no further FND symptoms. One of these patients has had a second round of tumour treatment without a recurrence of their FND. Three patients have persistent FND, but the FND symptoms were much less disabling than at presentation in two of those patients. One patient showed no improvement in their functional seizures.

#### Example case history: Acute FND delays glioma treatment

An adult patient developed daily attacks of non-pulsatile tinnitus. Otological and audiological investigations were normal. An MRI brain scan revealed a 15mm diameter, non-enhancing lesion in the left anterior insula (Figure 2). The lesion showed no restricted diffusion or increased perfusion. Interval scans showed a small increase in tumour size. The patient opted for surgical resection of the tumour. The patient needed to be woken during the operation to test their speech. While waiting for the operation, the patient developed a stutter. It was most pronounced when starting to speak. The patient was able to repeat some labial sounds e.g. m, m, m, but not p, p, p. However, they could not say words, such as mandarin, that began with a labial sound, m. There was no articulatory groping. Cranial nerve examination was normal. The MRI showed no new changes. An EEG recorded no epileptiform discharges. A diagnosis of a functional stutter was made. The patient was referred to speech and language therapists and neuropsychiatry for treatment. The patient’s speech returned to normal over several weeks. The tumour surgery was delayed for five months, but was performed successfully with intraoperative speech testing. The neuropathalogical tumour diagnosis was Astrocytoma IDH1 mutant, CNS WHO grade 2. The patient made a good recovery from the tumour surgery and returned to work.

**Figure 2.**
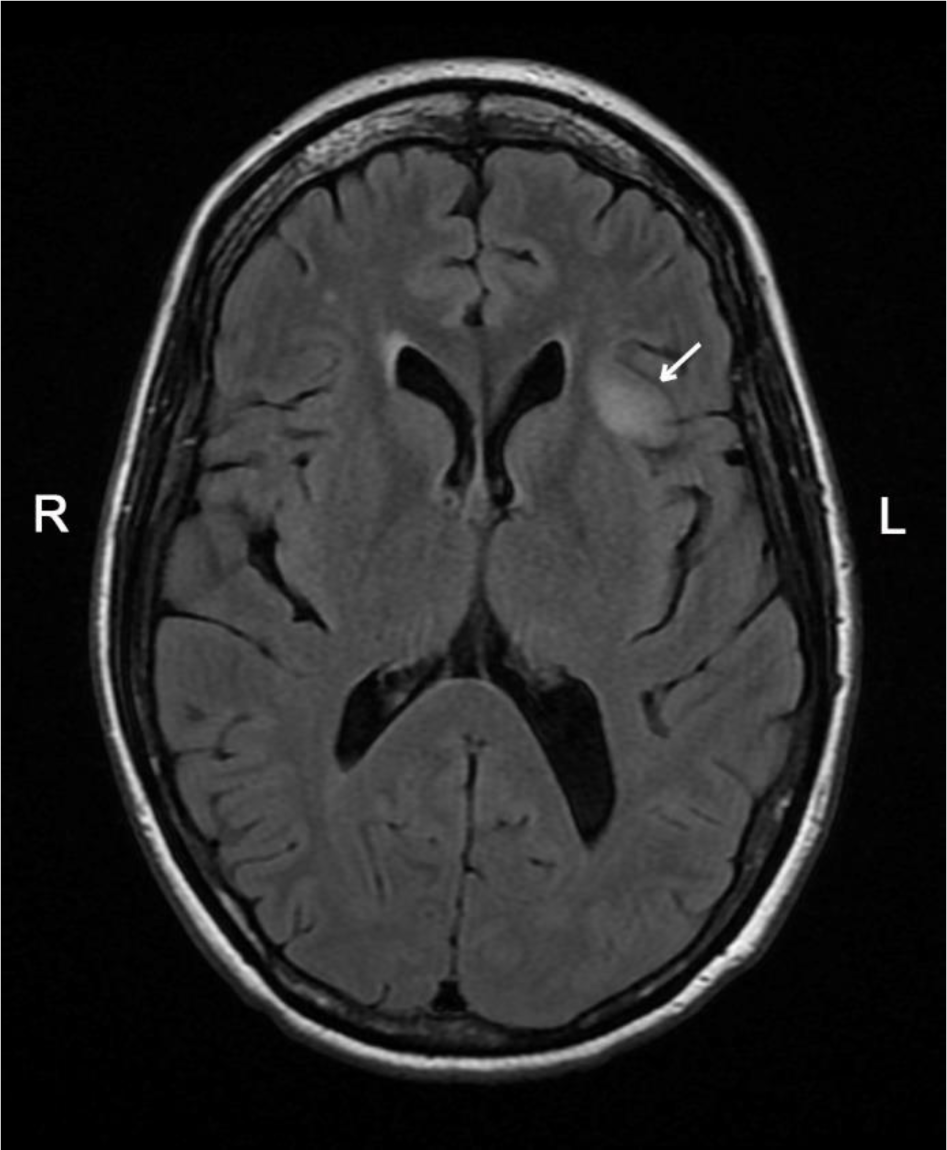
Low-grade glioma imaged with an axial fluid attenuated inversion recovery (FLAIR) MRI scan. White arrow points at the low-grade glioma (high FLAIR signal) in the left anterior insula. L, left; R, right.

### Systematic review

We identified ten papers reporting a total of 37 patients that met our review criteria (Figure 3). Four papers were case series and seven papers reported a single case (Table 2). There were no systematic reviews, clinical trials, intervention studies or case control studies of FND in neuro-oncology patients. The four case-series that we found were all retrospective studies of patients who developed functional seizures (Table 2).^18-21^ The case reports comprised three patients with functional seizures^22-24^ and four patients with motor symptoms (slurred speech and ataxia^25^; leg paresis^26^; torticollis^27^; functional gait disorder^28^).

**Figure 3.**
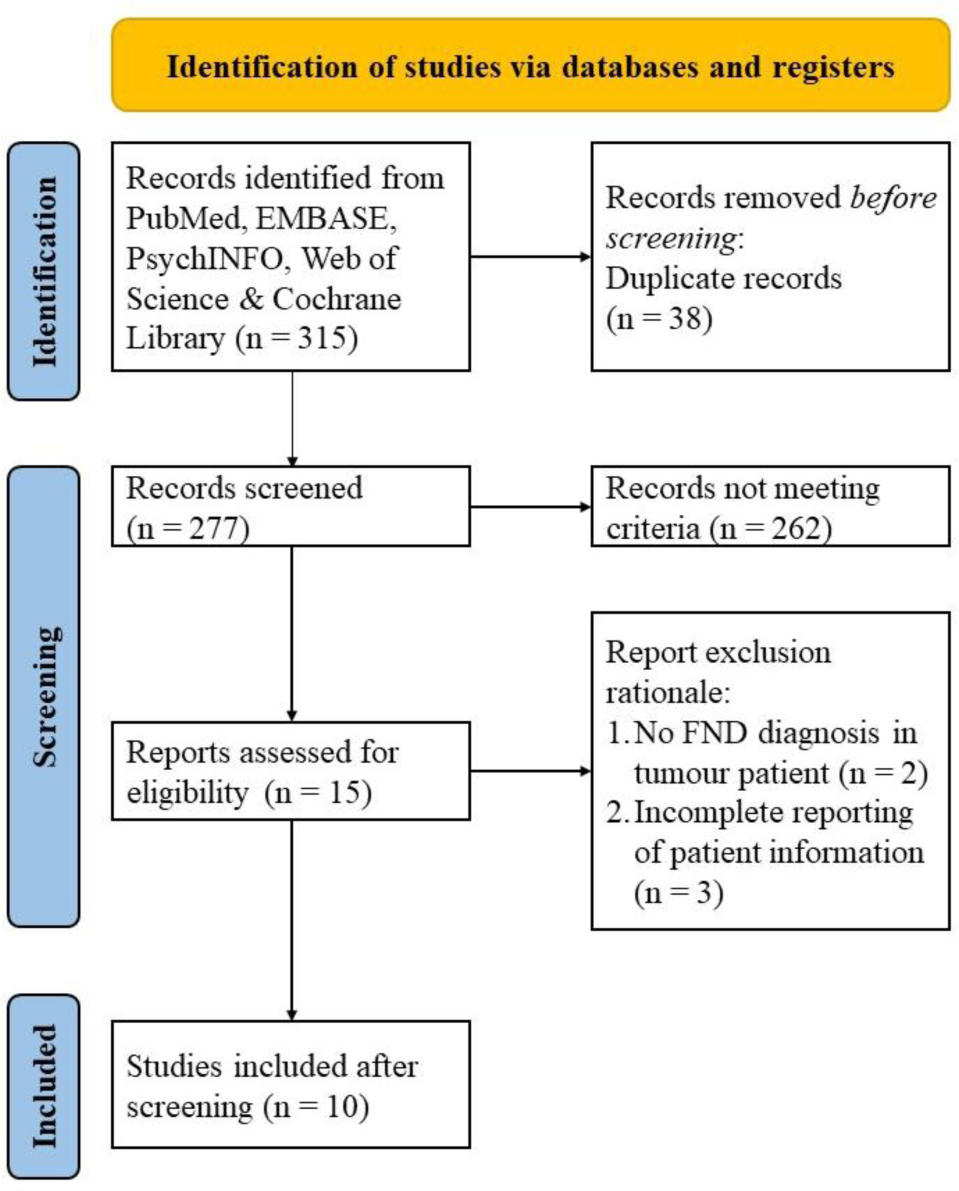
PRISMA flow diagram for systematic review. PRISMA, preferred reporting items for systematic reviews and meta-analyses.

**Table 2:**
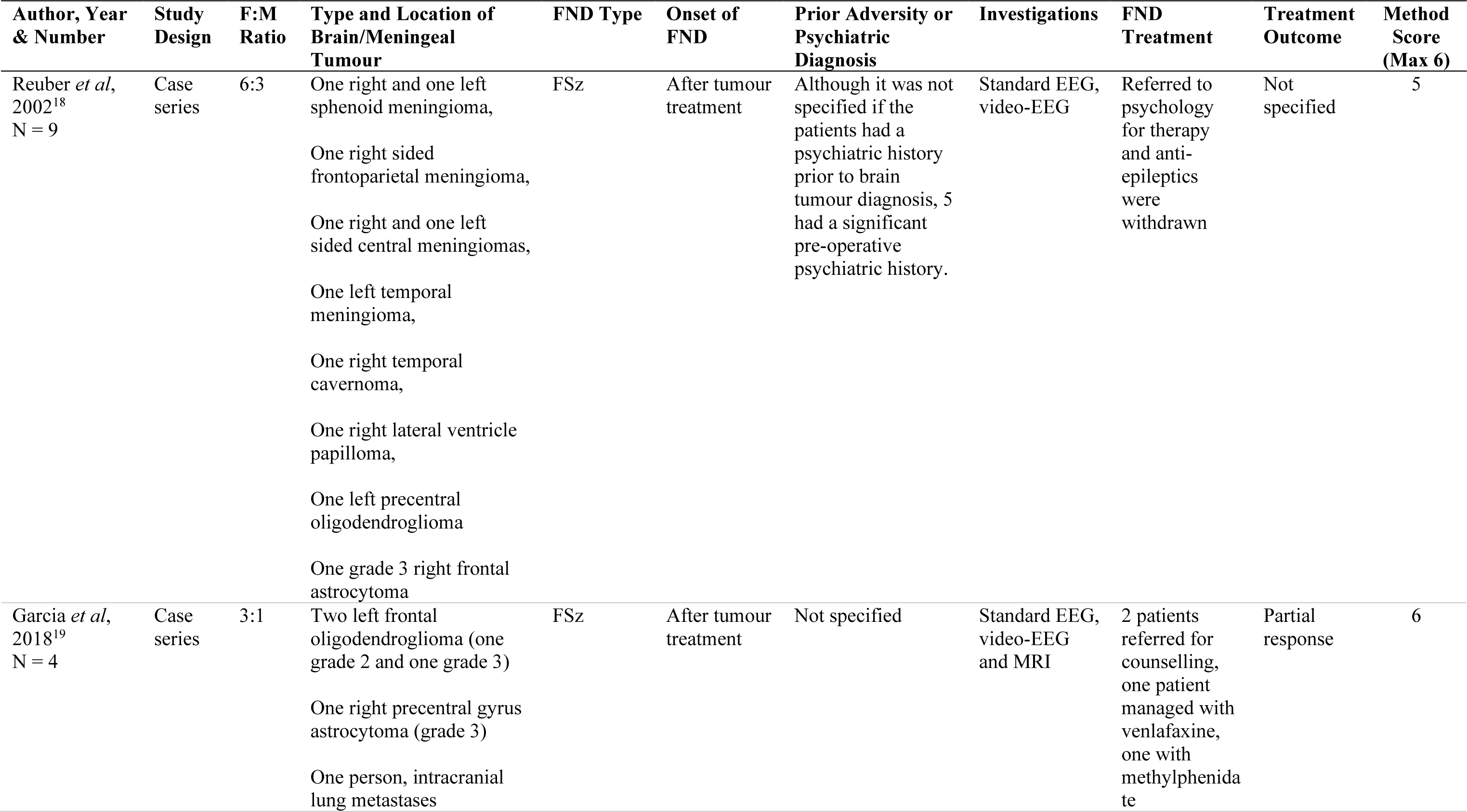
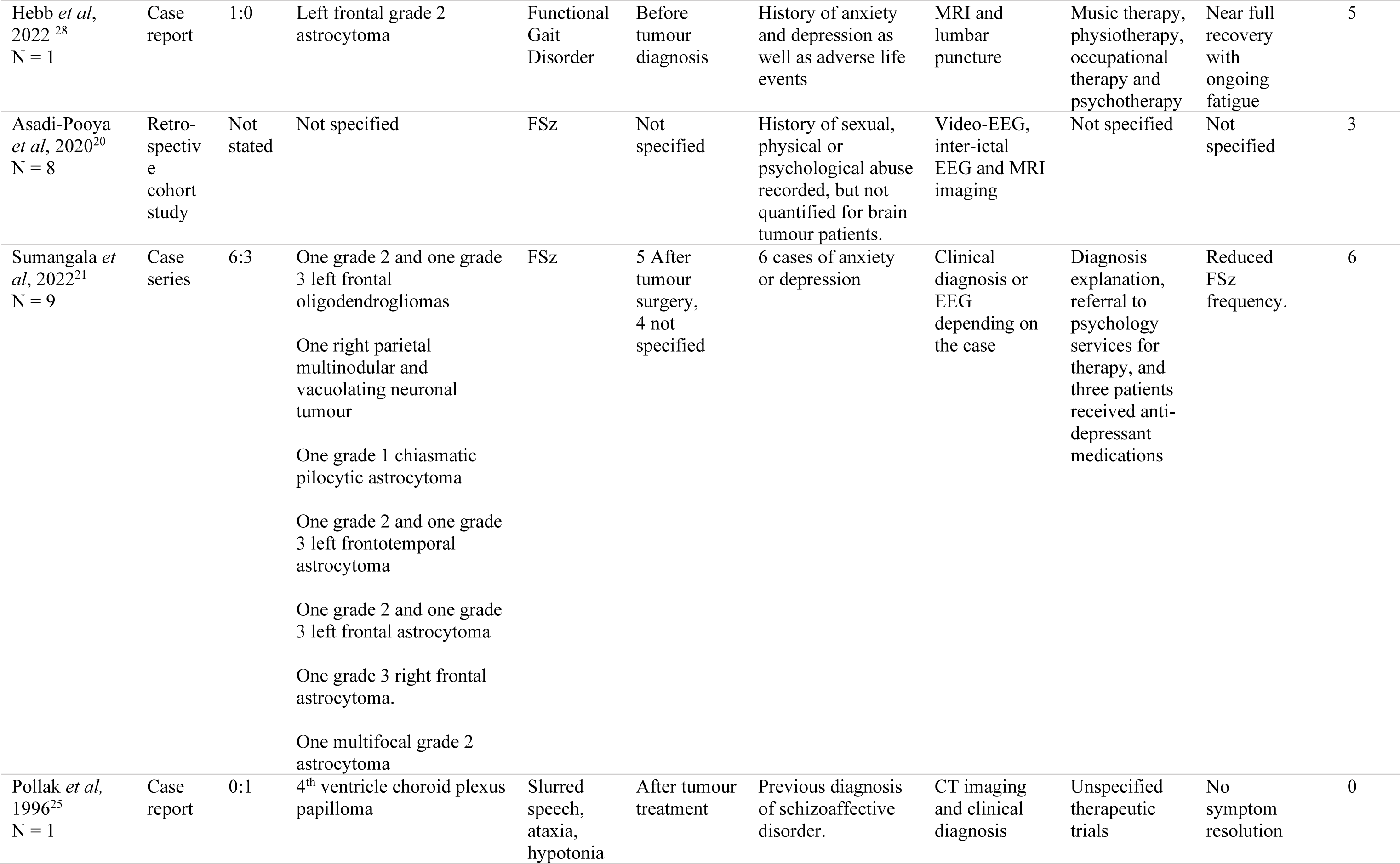
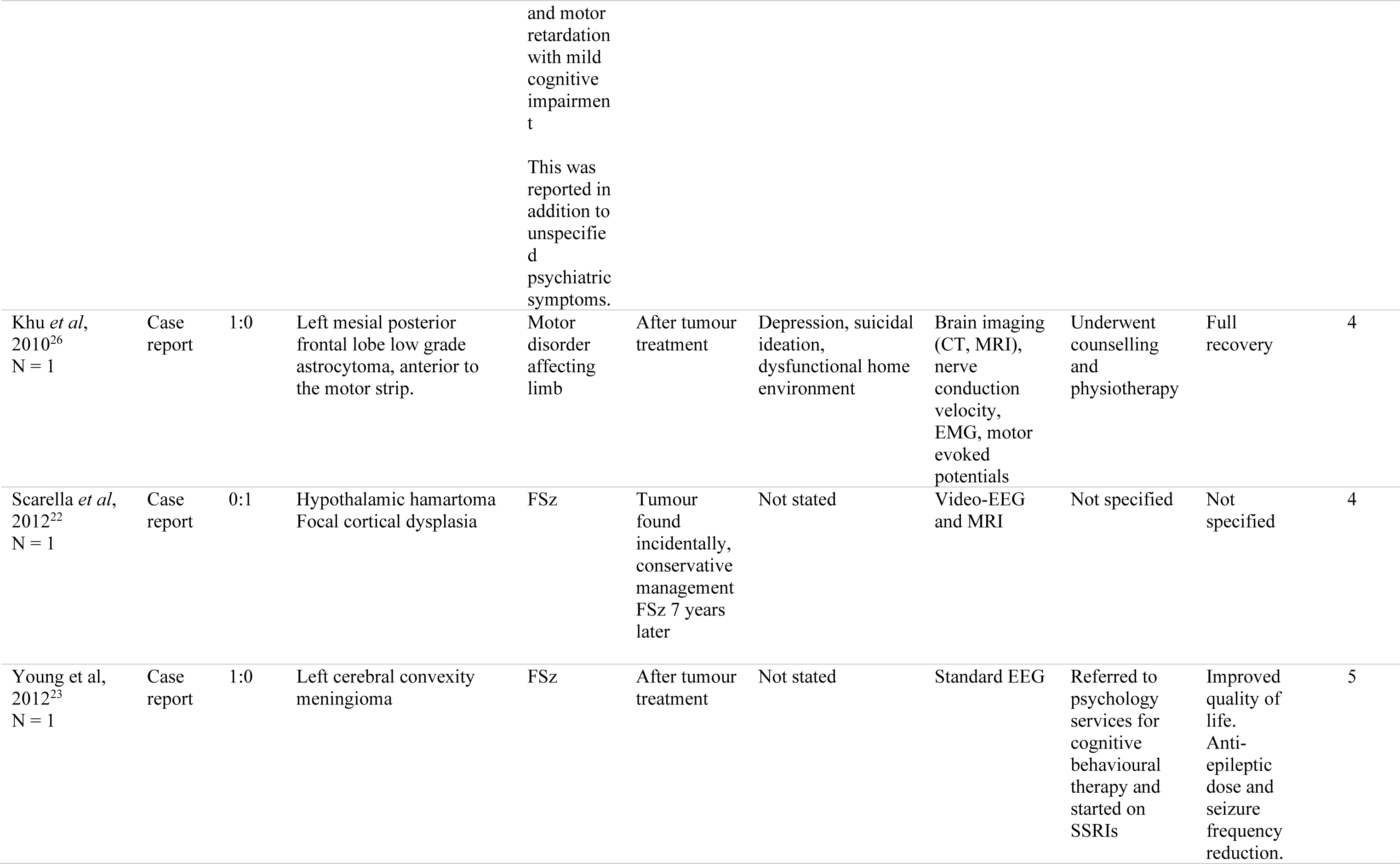
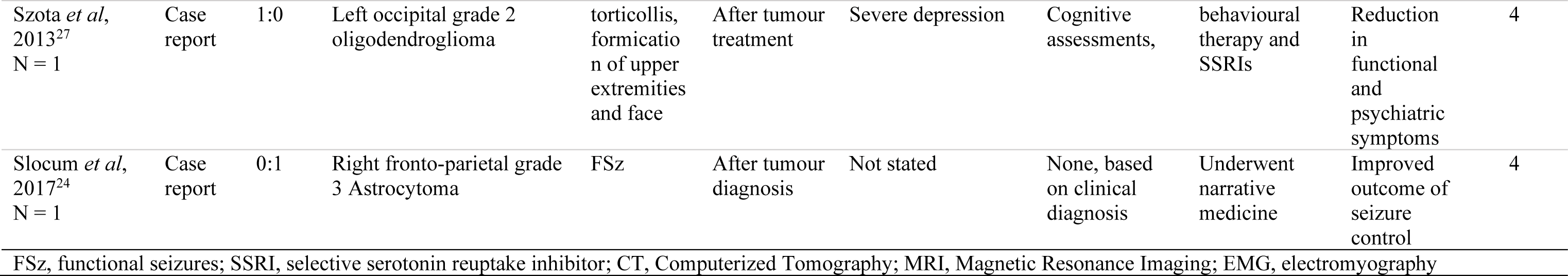
Papers reporting cases of functional neurological disorders in neuro-oncology patients.

36 patients in the systematic review developed FND symptoms after tumour treatment started or after a decision was made to manage the tumour conservatively (no surgery or radio-chemotherapy). Only one report described a patient with functional symptoms before their tumour diagnosis.^28^ The onset of the FND symptoms post treatment was recorded in three of the four case series and in three case reports. The median durations of tumour treatment to onset of FND symptoms in the case series were 4 months^21^, 12 months^18^, 13.5 months^19^ and in the case reports were <1 day^26^, <24 months^27^, >24 months^23^. The earliest onset of FND symptoms post treatment that we found in the literature was a case report of a patient that woke with functional limb paresis on the day of their brain tumour surgery.^26^ The FND symptoms of six patients in the systematic review started within one month of their tumour treatment beginning.

All patients had brain imaging, either computer tomography or MRI (Table 2). Patients with functional seizures also had an interictal EEG and/or video-telemetry. Limb weakness was investigated with nerve conduction studies, electromyography and motor evoked potentials.

FND were managed predominantly with psychological treatments including; counselling, behavioural therapy (unspecified), education to identify underlying stressors and other psychological therapy approaches, such as narrative medicine and music therapy (Table 2). Functional limb weakness was also treated with physiotherapy.

A psychiatric diagnosis had been made in 15 patients (depression and/or anxiety, 8; schizoaffective disorder, 1; unspecified, 5) either before the tumour diagnosis or during tumour treatment (Table 2). No psychiatric diagnosis was recorded for 22 patients. Serious adverse life events prior to the tumour diagnosis were not routinely reported. This information was recorded in two case reports and was asked about in one case series, but no breakdown was given for the brain tumour patients (Table 2).

The type of tumour and its location was specified in 29 patients. The predominant tumour types were lower-grade glioma (n = 17 patients) and meningiomas (n = 6 patients). The remaining 6 patients had five different types of tumour including one case of brain metastasis (Table 2). The tumour was located in the left hemisphere in 18 patients, right hemisphere in 7 patients and in the midline in 4 patients.

## DISCUSSION

Our case series focused on people with brain or meningeal tumour who developed a comorbid acute FND that negatively impacted their tumour treatment and required expedited investigations. Acute FND not only cause distressing symptoms in their own right, but have wider consequences for neuro-oncology patients. We found that FND could present acutely in neuro-oncology patients at any stage of the tumour illness. The symptoms of acute FND predominantly comprised functional seizures, functional hemiparesis, and functional speech disorders. A failure to diagnose and manage FND rapidly can result in inappropriate medical interventions and delays to tumour management. This is a problem when tumour treatment must occur within specific time limits. Finally, acute FND symptoms often became chronic.

Acute FND could occur at any stage of the tumour illness, but were more common in the interval that straddled diagnostic tumour investigations through to the period just after tumour treatment had started. Stress is a risk factor for FND^16 17^ and may be a contributing factor as neuro-oncology patients suffer acute stress during their tumour investigations and treatment.^29^

We found that acute FND could lead to unnecessary interventions, such as thrombolysis, and inappropriate treatment comprising overuse of anticonvulsant medication. Similar issues have been reported in people with acute FND that do not have tumours.^10 11^ A correct diagnosis of functional seizures enabled us to reduce or stop anticonvulsants in 50% of patients presenting with functional seizures.

Acute FND symptoms persisted for longer than 6 months in most of our patients. The prognosis of FND symptoms is mixed with some patients doing well^30 31^ and others poorly.^32^ Neuro-oncology patients often undergo multiple rounds of tumour treatment and, hence, remain at high risk of persistent or recurrent FND symptoms. Treatment that puts FND symptom into remission may prevent symptom recurrence during future treatment cycles: this was true for one of our patients. More work is needed to delineate the longer-term effects of acute FND on the quality of life of neuro-oncology patients.

People with lower-grade brain tumours and meningiomas are more at risk of their tumour treatment being affected. A contributing factor is that more aggressive tumours, such as glioblastoma, usually have a shorter interval between diagnosis and treatment compared with lower-grade gliomas and meningiomas. A larger study would be needed to establish whether brain tumours increase the risk of acute FND by disrupting specific neural circuits^33^ such as hypothesised modifications to the salience networks within the brain^34^ and/or through the stress associated with a neuro-oncology diagnosis.^7 16 17 29^

Our findings differ from the systematic review in the timing and frequency of FND types found in neuro-oncology patients. Firstly, all but one of the published cases of FND in brain tumour patients describe the FND symptoms developing after tumour treatment has started. In contrast, we found that acute FND symptoms can occur at any stage of the tumour-illness journey, most notably during the investigation period and in the run up to tumour treatment. Our findings suggest that acute FND prior to tumour treatment is an under-recognized problem.

The neurological presentations of FND that we describe are similar to those reported in the FND literature for non-tumour patients.^7 8^ Similarly, our finding that the semiology of functional seizures could be different from tumour-associated seizures has been described previously.^18 19 21^ However, the literature on FND in brain tumour patients is dominated by reports of functional seizures. This may be attributable to a selection bias in the literature for functional seizures as they are a form of FND that is well recognised due to differences in semiology and video EEG can confirm the diagnosis.

Diagnosing FND and explaining the condition carefully is sufficient in some patients to ameliorate the FND symptoms^30^ and enable tumour treatment to restart. This emphasizes the importance of early diagnosis and explanation of FND in management. However, other patients need more intensive intervention (Example case history) to minimize delays to tumour treatment and to prevent acute FND symptoms becoming chronic.

Treatment of comorbid FND has multiple goals including; alleviating FND symptoms to improve quality of life, prevent acute FND symptoms becoming chronic, minimize delays to neuro-oncology treatment and avoid iatrogenic harm from unnecessary treatments.

Neuro-oncology patients would benefit from a dedicated multi-disciplinary FND care pathway to expedite FND treatment and to prevent delays to tumour treatment. Neurologists are often the first point of contact when medical management fails.^35^ The care pathway needs to include neuropsychiatry^7^, physiotherapy ^36^, speech and language therapy^37^, psychological treatment^38^ and be integrated with existing cancer care services, including cancer psychologists and therapists.

In conclusion, neuro-oncology patients can develop acute FND that not only disable the patient, but also delay their tumour management. Greater awareness of this possibility will increase the speed with which the correct diagnosis is made and reduce the risk of inappropriate treatment. An expedited tumour-FND care pathway is needed to optimize treatment of the FND whilst minimizing consequences for brain tumour management.

## Data Availability

All data produced in the present work are contained in the manuscript

## Acknowledgements

We thank Dr Charmaine Toh for initial work on this project and Tim Nicholson and Mark Edwards for helpful comments on the manuscript. For the purposes of open access, the author has applied a Creative Commons Attribution (CC BY) licence to any Accepted Author Manuscript version arising from this submission.

## Contributors

GTF conceived the idea for the study. Analysis was performed by SI, DJ and GTF. SI, and GTF drafted the manuscript. All authors commented on the manuscript and gave approval for publication.

## Patient consent for publication

Patient consent was given to publish the example case history and MRI scan.

## Ethical Approval

Ethical approval for the study was obtained from the institutional review committee.

## Data Availability

All data relevant to this study are included in the article.

